# Pathway-based polygenic risk scores for schizophrenia and associations with clinical and neuroimaging phenotypes in UK Biobank

**DOI:** 10.1101/2022.07.12.22277553

**Authors:** Miruna C. Barbu, Gladi Thng, Mark J. Adams, Katie Marwick, Seth GN Grant, Andrew M. McIntosh, Stephen M. Lawrie, Heather C. Whalley

## Abstract

**Background:** Schizophrenia is a heritable psychiatric disorder with a polygenic architecture. Genome-wide association studies (GWAS) have reported an increasing number of risk-associated variants and polygenic risk scores (PRS) now explain 17% of the variance in the disorder. There exists substantial heterogeneity in the effect of these variants and aggregating them based on biologically relevant functions may provide mechanistic insight into the disorder.

**Methods:** Using the largest schizophrenia GWAS to date, we calculated PRS based on 5 gene-sets previously found to contribute to the pathophysiology of schizophrenia: the postsynaptic density of excitatory synapses, postsynaptic membrane, dendritic spine, axon, and histone H3-K4 methylation gene-sets. We associated each PRS, along with respective whole-genome PRS (excluding single nucleotide polymorphisms in each gene-set), with neuroimaging (N>29,000; cortical, subcortical, and white matter microstructure) and clinical (N>119,000; psychotic-like experiences including conspiracies, communications, voices, visions, and distress) variables in healthy subjects in UK Biobank.

**Results:** A number of clinical and neuroimaging variables were significantly associated with the axon gene-set (psychotic-like communications: β=0.0916, p_FDR_=0.04, parahippocampal gyrus volume: β=0.0156, p_FDR_=0.03, FA thalamic radiations: β=-0.014, p_FDR_=0.036, FA posterior thalamic radiations: β=-0.016, p_FDR_=0.048), postsynaptic density gene-set (distress due to psychotic-like experiences: β=0.0588, p_FDR_=0.02, global surface area: β=-0.012, p_FDR_=0.034, and cingulate lobe surface area: β=-0.014, p_FDR_=0.04), and histone gene-set (entorhinal surface area: β=-0.016, p_FDR_=0.035). In the associations above, whole-genome PRS were significantly associated with psychotic-like communications (β=0.2218, p_FDR_ =1.34×10^−7^), distress (β=0.1943, p_FDR_ =7.28×10^−16^), and FA thalamic radiations (β=-0.0143, p_FDR_=0.036). Permutation analysis carried out for these associations revealed that they were not due to chance.

**Conclusions:** Our results indicate that genetic variation in 3 gene-sets relevant to schizophrenia (axon, postsynaptic density, histone) may confer risk for the disorder through effects on a number of neuroimaging variables that have previously been implicated in schizophrenia. As neuroimaging associations were stronger for gene-set PRS than whole-genome PRS, findings here highlight that selection of biologically relevant variants may address the heterogeneity of the disorder by providing further mechanistic insight into schizophrenia.

## Introduction

Schizophrenia is a psychiatric disorder that is characterised by positive (hallucinations, delusions) and negative (anhedonia, avolition) symptoms, as well as marked cognitive impairment (1). Schizophrenia is thought to result from a complex combination of genetic and environmental factors, and its heritability is estimated at 80% (2).

Genome-wide association studies (GWAS) have reported increasing numbers of genomic loci associated with schizophrenia, lending support to the contribution of common genetic variants to the pathophysiology of schizophrenia (3,4). Genome-wide polygenic risk scores (PRS) calculated from GWAS explain approximately 17% (Nagelkerke’s R^2^) of the variance in schizophrenia (4). However, there exists substantial heterogeneity in the effect of risk variants, and genome-wide approaches may not be sufficient for patient stratification in downstream analyses. Mechanistic insight may be derived from GWASs, including the identification of biologically relevant gene sets in which risk variants are aggregated. For instance, The Network and Pathway Analysis Subgroup of the Psychiatric Genomics Consortium (PGC) investigated significant pathways across three adult psychiatric disorders: schizophrenia, major depressive disorder, and bipolar disorder. They identified a number of pathways specific to each disorder, but also common across the three, including histone methylation, immune, and neuronal signalling pathways (5).

Combining the predictive power of PRS with findings from pathway analysis by examining genetic variation within biologically relevant gene sets may address the inherent heterogeneity in schizophrenia and may provide additional mechanistic insight by detecting associations with biologically informative traits. This methodology has been applied to a number of relevant phenotypes to schizophrenia. For instance, Rampino et al. (2017) found that schizophrenia PRS calculated based on single nucleotide polymorphisms (SNPs) implicated in glutamatergic signalling were associated with attention, a cognitive process known to be impaired in schizophrenia (6). Yao et al. (2021) found that PRS calculated based on neural *microRNA-137 (MIR137)* explained a disproportionately larger schizophrenia risk variance than genomic control PRS, when accounting for gene-set size (∼2% *MIR137* N = ∼1,000 genes; ∼10%, whole-genome N= ∼20,000 genes) (7). It is therefore possible to interrogate genetic risk aggregated to biologically relevant gene-sets to gain insight into the association between aggregated genetic risk and traits of interest to the disorder.

Schizophrenia PRSs have also been investigated in relation to the organ of interest in the disorder, the brain. Previous evidence has shown associations with disruptions in white matter microstructure and global and regional brain volumes, although results are inconsistent (8,9). As such, it was suggested that perhaps biologically relevant gene sets may reveal stronger associations with neuroimaging phenotypes, as demonstrated recently (10). Grama et al. (2020) investigated whether behaviour- and neuronal-related gene-sets, previously found to be implicated in schizophrenia, are associated with subcortical volumes. They found that one gene-set, “abnormal behaviour”, was associated with right thalamic volume, and this association was robust across different p-value thresholds (10). This methodology can also be applied to other psychiatric disorders. For instance, PRS calculated for a previously established biological pathway (NETRIN1) in relation to major depressive disorder (MDD) were associated with relevant neuroimaging phenotypes, shedding light on links between biology and neuroimaging (11). This indicates that meaningful associations with traits of interest may be revealed when applying genomic methods addressing relevant parts of the genome.

Based on this evidence, we hypothesised that schizophrenia PRS aggregated in biologically relevant pathways that have previously been shown to play a role in schizophrenia would be associated with structural neuroimaging and clinical phenotypes in a general population adult sample. Identifying associations with specific neuroimaging phenotypes may provide an opportunity to disentangle the heterogeneity of the disorder, both in terms of genetic risk and inconsistent previous neuroimaging findings. We therefore selected 5 gene-sets, specifically: postsynaptic density (PSD), postsynaptic membrane (PSM), dendritic spine, axon, and histone H3-K4 methylation, previously identified by The Network and Pathway Analysis Subgroup of the PGC (5). These 5 cellular components and biological processes have previously been associated with schizophrenia in a number of other studies investigating both human and animal models (12–14). We calculated PRSs for each gene-set-specific set of SNPs and SNPs excluded from the gene-sets for paired comparisons (gene-set SNPs versus whole-genome (WG) minus gene-set SNPs PRS). We then tested their association with brain structural phenotypes (cortical volumes, thickness, and surface area; white matter microstructure as indexed by fractional anisotropy (FA) and mean diffusivity (MD); and subcortical volumes) and psychotic-like experiences (PLEs, Mental Health Questionnaire (MHQ)) in UK Biobank (UKB). We utilised the most up-to-date genetic, mental health, and imaging data. We hypothesized distinctive roles in the pathophysiology of schizophrenia for the different biologically relevant pathways tested, after adjustment for whole-genome PRSs, highlighting important mechanistic processes underlying the different phenotypes associated with schizophrenia. We expect this methodology to partly address the heterogeneity in schizophrenia through the identification of biologically relevant mechanisms.

## Methods and Materials

### Study population

UKB comprises 502,411 community-dwelling individuals whose information was recruited between 2006 and 2010 in the United Kingdom (https://biobank.ctsu.ox.ac.uk/crystal/field.cgi?id=200) (15). UKB received ethical approval from the research ethics committee (REC reference 11/NW/0382), and this study has been approved by the UKB Access Committee (Project No. 4844 and 16124). Written informed consent was obtained from all participants. The current study was conducted using the latest release of UKB neuroimaging data (N_cortical_=29,791, N_subcortical_=29,536, N_white matter_=27,917) as well as N=119,947 with MHQ data. We excluded individuals with a diagnosis of schizophrenia as indicated by ICD-10 (F20, https://biobank.ndph.ox.ac.uk/showcase/field.cgi?id=41270) due to the small proportion of diagnosed individuals (N=989) and due to the fact that our analyses focussed on associations in the general population.

### Gene-set selection

The top 5 gene-sets associated with schizophrenia were selected from The Network and Pathway Analysis Subgroup of the Psychiatric Genomics Consortium (5) and identified on Gene Ontology (GO) by searching for each pathway’s GO identifier in the discovery manuscript above (5,16): postsynaptic density (GO:0014069), postsynaptic membrane (GO:0045211), dendritic spine (GO:0043197), axon (GO:0030424), and histone H3-K4 methylation (GO:0051568). The gene-sets were selected based on their robust association with schizophrenia in this, and previous studies (5,13,17,18). Further details on each gene-set are included in the Supplementary Materials and Supplementary Excel File 1.

### Genotyping, SNP annotation and PRS profiling

Genotyping of 488,000 blood samples from UKB participants was carried out using the UK BiLEVE (https://biobank.ctsu.ox.ac.uk/crystal/refer.cgi%3fid%3d149600) or UKB Axiom (https://biobank.ctsu.ox.ac.uk/crystal/refer.cgi%3fid%3d149601) arrays (19). Further information on genotyping procedures and quality control are provided in https://biobank.ctsu.ox.ac.uk/crystal/crystal/docs/genotyping_qc.pdf and in Bycroft et al. (19). Genetic data from both the base and target datasets were annotated in reference to human genome build 19.

We used SNPs from the largest available GWAS of schizophrenia (N=320,404, N=76,755 cases), carried out by Trubetskoy et al. (4) as the base dataset. Here, they identified schizophrenia associations with common variants at 287 distinct loci, and PRSs explained approximately 17% of the variance in schizophrenia in a European ancestry target sample (4). The GWAS sample utilised here did not include any individuals in UKB. The following quality check (QC) was performed based on the protocol by Choi et al. ((20), https://choishingwan.github.io/PRS-Tutorial/base/): removing SNPs with a minor allele frequency (MAF) < 0.01, and imputation information score < 0.8; removing duplicated and ambiguous SNPs; and ensuring that there were no overlapping samples with the target dataset (UKB). This resulted in a final dataset containing N=5,899,138 SNPs. QC in UKB was performed based on the same protocol (20) (https://choishingwan.github.io/PRS-Tutorial/target/): removing SNPs with a MAF < 0.01, Hardy-Weinberg Equilibrium (HWE) < 1×10^−6^, SNPs that are missing in a high proportion of participants (<0.02), highly-correlated SNPs (r^2^=0.25); and removing individuals that have a high rate of genotype missingness (0.02) and that contain high heterozygosity rates (> 3 SDs from the mean). Additionally, we ensured that our sample consisted of unrelated, white British participants with no overlap with PGC (21). The final genetic sample consisted of N=365,125 participants and N=5,974,990 SNPs, and this was reduced further when combining with clinical and imaging samples (see below).

ANNOVAR was utilised to annotate SNPs in the base dataset, if they were located between 20 kb upstream or downstream of the transcription start and end sites, respectively (22). Following functional annotation, SNPs located within each gene-set were extracted. See Supplementary Table 1 and Supplementary Excel File 1 for the number of genes and SNPs within each gene-set. These lists were used as input for the creation of gene-set PRSs as described below, as well as for creation of whole-genome PRS exclusive of each gene-set (i.e. each gene-set SNP list was input as an exclusion flag to create whole-genome PRS that did not include the specific gene-sets).

Gene-set PRSs for each individual in UKB were computed using PRSice (23) at 5 p-value thresholds (0.01, 0.05, 0.1, 0.5, and 1) by summing the number of risk alleles weighted by the strength of association with schizophrenia in the base dataset. Each gene-set PRS had its respective whole-genome PRS (WG PRS). PRSs were created with clumping-based pruning of SNPs in linkage disequilibrium (r^2^=0.25, 500-kb window). SNPs within the extended MHC locus (chr6:25 Mb–35Mb) were excluded due to high LD in the region. The primary analysis in this manuscript comprises SNPs that met a significance level of 0.1 (4). Analyses involving all other thresholds are included in the Supplementary Excel File 2 (Tables 1-7).

**Table 1.** Demographic characteristics for the clinical samples.

|  | **MHQ** |
| --- | --- |
| Age mean (SD) | 56.09 (7.69) |
| Sex, F (%) | 67,432 (56%) |
| MHQ – heard unreal voice (Yes, %) | 1,918 (1.6%) |
| MHQ – seen unreal vision<br>(Yes, %) | 3,696 (3%) |
| MHQ – believed in unreal conspiracy (Yes, %) | 724 (0.60%) |
| MHQ – believed in unreal communications (Yes, %) | 652 (0.54%) |
| Distressing PLEs (Yes, %) | 2,046 (1.76%) |

### Phenotypes

#### Psychotic-like experiences

A Mental Health Questionnaire (MHQ) was sent to participants who provided an e-mail address from July 2016 to July 2017 (approximately 157,000 individuals). There were four unusual and psychotic experiences items included in the MHQ. Here, we utilised dichotomised responses on the following questions: “Did you ever believe that there was an unjust plot going on to harm you or to have people follow you, and which your family and friends did not believe existed?”, “Did you ever believe that a strange force was trying to communicate directly with you by sending special signs or signals that you could understand but that no one else could understand (for example through the radio or television)?”, “Did you ever hear things that other people said did not exist, like strange voices coming from inside your head talking to you or about you, or voices coming out of the air when there was no one around?”, and “Did you ever see something that wasn’t really there that other people could not see?”. In addition, we also examined distress by selecting individuals who reported PLEs as distressing (as opposed to neutral or positive) and those who reported no PLE to determine associations with experiencing stress (24–26). The four lifetime PLEs will be referred to as “conspiracies”, “communications”, “voices”, and “visions”, based on the clinical significance of each PLE, and distressing PLEs will be referred to as “distress”. Frequencies for all items are noted in Table 1.

#### Neuroimaging phenotypes

A brain MRI scan was conducted for a subset of participants (27), and imaging-derived phenotypes (IDPs) were used in this study. T1 weighted and diffusion (DTI) MRI images were acquired using a Siemens Skyra 3 T scanner with a standard Siemens 32-channel RF receive head coil. Magnetic resonance imaging (MRI) acquisition, pre-processing, and QC for all structural phenotypes were performed utilising standardised protocols in UKB and are described in detail elsewhere (https://biobank.ctsu.ox.ac.uk/crystal/crystal/docs/brain_mri.pdf) (27,28). Exclusion criteria comprised removal of scans with severe normalization problems by the UKB (28). Individuals whose global values were >3 SDs from the sample mean were also excluded (range N excluded based on imaging modality=105-232 participants, distribution plots created using “ggplot” in R are shown in Supplementary Figure 1) (29,30).

A list of all neuroimaging phenotypes investigated is included in Supplementary Table 2. Briefly, parcellation of white matter tracts was conducted using AutoPtx (27) and resulted in 15 white matter tracts (3 unilateral). Two DTI scalars, FA and MD, were utilised here (https://biobank.ndph.ox.ac.uk/showcase/label.cgi?id=135). Cortical regions of interest (ROIs) were identified using Desikan-Killiany-Tourville parcellation in FreeSurfer, and resulted in 31 cortical structures per hemisphere (31,32) (https://biobank.ndph.ox.ac.uk/showcase/label.cgi?id=196). Here, we investigated cortical thickness (CT), surface area (SA), and cortical volume (CV). Finally, 8 subcortical grey matter ROIs per hemisphere were identified (33). Sample size and descriptive statistics for all neuroimaging phenotypes are in Table 2.

**Table 2.** Demographic characteristics for the neuroimaging samples. N is different for each variable as outlier exclusion (3 SDs from mean) was applied individually to each phenotype.

|  | Cortical regions | Subcortical volumes | White matter microstructure |
| --- | --- | --- | --- |
| Age mean (SD) | 63.71 (7.48) |  |  |
| Sex, F (%) | 15,666 (53%) | 15,557 (53%) | 14,745 (53%) |
| Scan site |  |  |  |
| Cheadle | 24,910 |  |  |
| Reading | 5,064 |  |  |
| Newcastle | 9,968 |  |  |

In addition to the specific regions and tracts mentioned above, we also derived general and regional measures for white matter microstructure and cortical phenotypes. For white matter microstructure, we derived global and regional (association and projection fibres, thalamic radiations) measures of FA and MD by conducting principal component analysis (PCA) on sets of tracts and extracting scores of the first unrotated principal component. For cortical ROIs, we derived global and lobar measures by summing up all (global) or lobar-specific ROIs, as in previous studies (34,35).

### Statistical analysis

All analyses were conducted using R (version 4.1.0) in a Linux environment. We used linear mixed-effects models (function “lme” in package “nlme”) for all bilateral neuroimaging phenotypes, with age, age^2^, sex, 15 genetic principal components (PC), scan site, three magnetic resonance imaging head position coordinates (lateral brain position X, https://biobank.ctsu.ox.ac.uk/crystal/field.cgi?id=25756, transverse brain position Y, https://biobank.ctsu.ox.ac.uk/crystal/field.cgi?id=25757, longitudinal brain position Zhttps://biobank.ctsu.ox.ac.uk/crystal/field.cgi?id=25757), and genotype array set as covariates. Hemisphere was included as a within-subject covariate in all mixed-effects models. Intracranial volume (ICV) was included as a covariate for all grey matter phenotypes. We used general linear models (function “lm”) for unilateral, regional, and global neuroimaging phenotypes, using the same covariates as above and without hemisphere included as a separate term. Finally, we used logistic regression for all dichotomised PLEs, with age, sex, 15 genetic PCs, and genotype array set as covariates. All models included gene-set PRS and each gene-set’s respective whole-genome PRS as predictor variables. False Discovery Rate (FDR) was used to correct for multiple testing and was applied separately for each neuroimaging and clinical phenotype and across all p-value thresholds (36). Effect sizes from linear models were standardised throughout.

In order to establish that the effect of gene-set PRS on neuroimaging and clinical phenotypes was not due to chance (as the SNP set sizes in all gene-sets were much smaller compared to the whole genome (Supplementary Table 1)), a permutation analysis was carried out using a method developed by Cabrera et al. (37) for all significant associations. First, for significant gene-set associations, circular genomic permutation was applied, by placing all SNPs in the genome (excluding those pertaining to each of the gene-sets demonstrating significance) in a circular genome, based on their location. One thousand SNP lists with the same set size as each of these gene-sets were then permuted. Then, we created 1,000 PRSs based on these SNP lists, that were then included in regression models for the phenotypes that were originally identified to be significantly associated with the gene-set PRSs. Significance was then determined by comparing t-values from the real associations with those derived from the permuted regression models. The permutation p-value was calculated by observing the position of the real t-values in the list of permuted t-values and dividing the position by the number of permutations (N=1,000).

## Results

All results presented below concern PRS at p-value threshold < 0.1. Descriptive statistics for clinical and neuroimaging phenotypes are noted in tables 1 and 2 below, respectively. Analyses involving all other thresholds are included in the Supplementary Excel File 2 (Tables 1-7).

### Demographic characteristics

*Table 1 and 2 go here*.

### Associations with psychotic-like experiences

Gene-set-specific significant findings and corresponding WG results are noted in Figure 1 below. The axon gene-set PRS was significantly associated with communications MHQ item (gene-set PRS β=0.0916, p_FTR_ =0.04; whole-genome PRS β=0.2218, p_FTR_=1.34×10^−7^), while the PSD gene-set PRS was significantly associated with distress associated with PLEs (gene-set PRS β=0.0588, p_FTR_ =0.02; whole-genome PRS β=0.1943, p_FTR_ =7.28×10^−16^). Whole-genome PRS were associated with all four MHQ items, as well as distress, and effect sizes were of greater magnitude than associations with gene-set PRS (see Supplementary Excel File 2, Table 1). Effect sizes for gene-set-specific PRS ranged from 1×10^−4^ to 0.092 and for whole-genome PRS from 0.087 to 0.300. Results concerning other MHQ items at other p-values thresholds are in the Supplementary Excel File 2 (Table 1).

**Figure 1.**
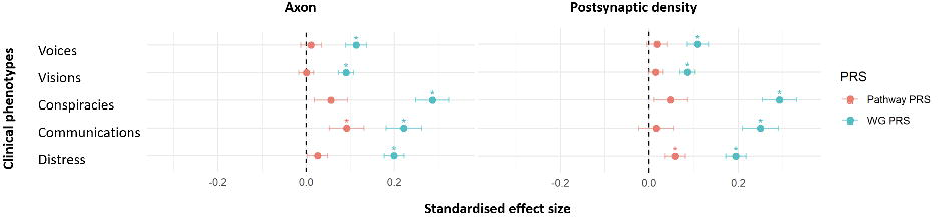
Significant associations between the axon and postsynaptic density gene-set PRSs and clinical phenotypes. X-axis indicates standardised effect sizes; *=significant associations after correction for multiple comparisons; WG = whole-genome.

### Associations with neuroimaging phenotypes

#### Gene-set PRS associations

The axon gene-set PRS were associated with parahippocampal gyrus volume (gene-set PRS β=0.0156, p_FDR_=0.03; whole-genome PRS β=-0.003, p_FDR_=0.833), FA thalamic radiations tract category (gene-set PRS β=-0.014, p_FDR_=0.036; whole-genome PRS β=-0.0143, p_FDR_=0.036), and FA posterior thalamic radiations (gene-set PRS β=-0.016, p_FDR_=0.048; whole-genome PRS β=-0.011, p_FDR_=0.126).

The PSD gene-set PRS were associated with global surface area (gene-set PRS β=-0.012, p_FDR_=0.034; whole-genome PRS β=-0.003, p_FDR_=0.517) and cingulate lobe surface area (gene-set PRS β=-0.014, p_FTR_ =0.04; whole-genome PRS β=1×10^−4^, p_FTR_ =0.977).

Finally, the histone H3-K4 methylation gene-set PRS were associated with entorhinal surface area (gene-set PRS β=-0.016, p_FDR_=0.035; whole-genome PRS β=0.01, p_FDR_=0.164).

Neuroimaging phenotypes associated with gene-set PRSs are indicated in Figures 2 and 3 below.

**Figure 2.**
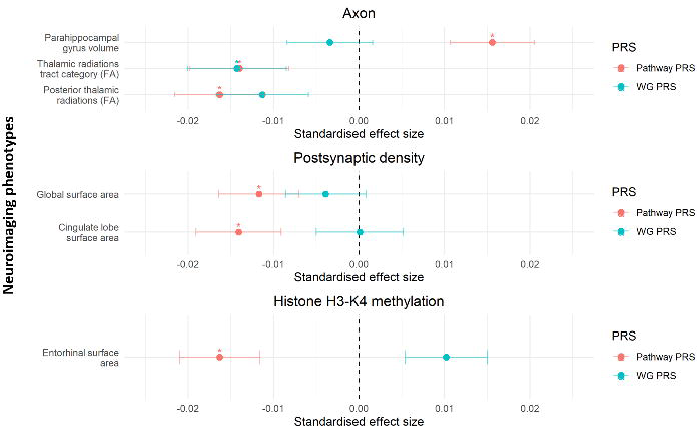
Significant associations between the axon, postsynaptic density, and histone H3-K4 methylation gene-set PRSs and neuroimaging phenotypes. X-axis indicates standardised effect sizes; *=significant associations after correction for multiple comparisons; WG = whole-genome.

**Figure 3.**
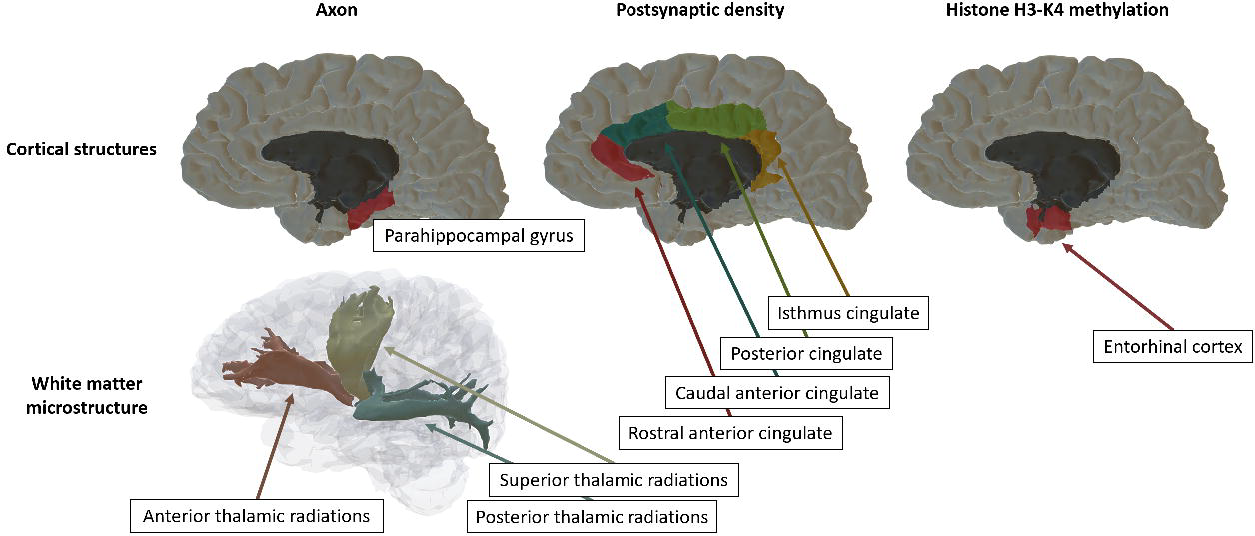
Cortical and white matter microstructure phenotypes that were associated with the axon, PSD, and histone gene-set PRSs. The thalamic radiations tract category, comprised of the anterior, superior, and posterior thalamic radiations, was associated with the axon gene-set PRS; the cingulate lobe, comprised of caudal anterior, rostral anterior, posterior and isthmus cingulate, was associated with PSD gene-set PRSs. Global surface area (i.e. the entire brain’s surface area) was also associated with PSD but is not indicated in the graph above.

#### Whole-genome PRS associations

Whole-genome PRS, irrespective of which SNPs were removed prior to calculating PRSs, were associated with a number of neuroimaging phenotypes, indicated below and in the Supplementary Excel File 2 (Tables 2-7). All standardised beta values are absolute, and ranges are not reported if they are rounded to the same number. Results presented below are consistent across whole-genome PRS that had different gene-set SNPs excluded.

FA: global FA (β range=0.016-0.019, p_FDR_ range=0.004-0.03), association fibres (β range=0.018-0.021, p_FDR_ range=0.003-0.019), thalamic radiations (β range=0.014-0.018, p_FDR_ range=0.006-0.03), cingulate gyrus (β range=0.014-0.015, p_FDR_ range=0.014-0.034), anterior (β range=0.016-0.017, p_FDR_ range=0.019-0.038) and posterior thalamic radiations (β range=0.015-0.016, p_FDR_ range=0.025-0.03), and inferior longitudinal (β range=0.015-0.017, p_FDR_ range=0.015-0.043) and inferior fronto-occipital fasciculi (β range=0.015-0.016, p_FDR_ range=0.024-0.035).

MD: global MD (β range=0.015-0.017, p_FDR_ range=0.011-0.04), association fibres (β range=0.013-0.014, p_FDR_ range=0.034-0.042), thalamic radiations (β range=0.014-0.016, p_FDR_ range=0.011-0.04), projection fibres (β range=0.014, p_FDR_ range=0.04), corticospinal tract (β range=0.02-0.022, p_FDR_ range=0.005-0.023), anterior (β range=0.015, p_FDR_ range=0.037), superior, (β range=0.014, p_FDR_ range=0.043), and posterior thalamic radiations (β range=0.013, p_FDR_ range=0.04), inferior longitudinal fasiculus (β range=0.015, p_FDR_ range=0.04), and cingulate gyrus (β range=0.015, p_FDR_ range=0.04).

Thalamus (β range=0.019-0.023, p_FTR_ range=3.6×10^−5^-0.0009) and accumbens (β range=0.013-0.015, p_FDR_ range=0.005-0.032) were also significantly associated with whole-genome PRS after multiple comparison corrections. Finally, significant cortical regions included the volume of the medial orbitofrontal cortex (β range=0.016-0.019, p_FDR_ range=0.003-0.027) and superior temporal gyrus (β range=0.018-0.02, p_FTR_ range=8.3×10^−4^-0.009), and the surface area of superior temporal gyrus (β range=0.015-0.016, p_FDR_ range=0.022-0.04).

### Permutation analysis

For those gene-set PRSs that were significantly associated with PLEs, white matter microstructure or cortical regions (axon: psychotic-like communications, volume of parahippocampal gyrus, thalamic radiations tract category, posterior thalamic radiations; PSD: distress, surface area of the cingulate lobe and global surface area; histone H3-K4 methylation: entorhinal cortex surface area), we performed circular genomic permutation analysis. We found that the significant associations with the gene-set PRSs in these clinical and neuroimaging phenotypes were not due to chance, based on a permutation p-value computed by comparing t-values from the real associations with t-values from the permuted associations (Table 3).

**Table 3.** Permutation results for gene-set PRS where significant associations were identified (PRS p-value threshold 0.1).

| Gene-set PRS | Phenotype | Gene-set PRS $\beta$ | Gene-set PRS t-value | Gene-set PRS calculated p-value |
| --- | --- | --- | --- | --- |
| Axon | Communications | 0.0916 | 2.306 | 0.026 |
|  | Parahippocampal gyrus volume | 0.0156 | 3.169 | 0.003 |
|  | Thalamic radiations (FA) | -0.014 | -2.44 | 0.01 |
|  | Posterior thalamic radiations (FA) | -0.016 | -3.055 | 0.004 |
| Postsynaptic density | Distress | 0.0588 | 2.618 | 0.01 |
|  | Cingulate lobe surface area | -0.014 | -2.812 | 0.012 |
|  | Global surface area | -0.012 | -2.498 | 0.024 |
| Histone H3-K4 methylation | Entorhinal cortex | -0.016 | -3.454 | 0.002 |

## Discussion

In this study, we aimed to investigate the association between biologically relevant pathway PRSs and clinical and neuroimaging phenotypes, after adjustment for respective whole-genome PRSs in N = ∼ 29,000 (neuroimaging phenotypes) and N = 119,947 (clinical phenotypes). We calculated PRSs based on the largest, most recent schizophrenia GWAS to date (4). We found significant associations for the axon, PSD, and histone H3-K4 methylation gene-sets with a number of cortical regions and white matter tracts, as well as with PLEs, specifically psychotic-like communications and distress associated with having a PLE. Associations with PLEs were stronger for whole-genome PRS, while associations with neuroimaging variables were stronger for gene-set PRS.

The three gene-sets with identified significant associations here have previously been involved in schizophrenia. The Network and Pathway Analysis Subgroup of the Psychiatric Genomics Consortium (5) identified the three pathways to be implicated in schizophrenia through robust computational analyses, by aggregating gene-sets from a large number of databases. The most significant genetic contribution to schizophrenia was found in genes encoding proteins located in excitatory synapses and in particular, the proteome of the PSD, followed by histone methylation-related processes. The PSD of excitatory synapses comprises protein complexes that assemble glutamate-sensitive neurotransmitter receptors to intracellular proteins (38,39). Postsynaptic networks have long been implicated in schizophrenia (40–42) as they play a role in cognition and synaptic plasticity, features that are known to be disrupted in schizophrenia (13,43). In addition, both post-synaptic and pre-synaptic networks were uncovered through gene prioritisation in the latest schizophrenia GWAS, highlighting the consistency of associations with this pathway over time as well as further rationale to investigate the pathway in this study (4).

Here, we identified associations between the PSD gene-set and decreased global and cingulate lobe surface area, as well as higher distress associated with PLEs. The cingulate lobe is part of the limbic system and comprises four cortical regions, specifically rostral, caudal, posterior, and isthmus cingulate. The region is involved in behaviour and emotion regulation and cognitive processes including memory, attention, and motivation, which are disrupted in schizophrenia (44). Volumes in this region were reduced in previous studies investigating their association with schizophrenia, and a systematic review concluded that hypoactivity of the cingulate cortex underlies the manifestation of negative symptoms in many patients, although the studies analysed provided inconsistent results (44,45). Interestingly, whole-genome PRS did not show associations with cingulate or global surface area in previous studies (46,47). Associations identified here indicate that narrowing the genome to a biologically relevant gene-set may reveal associations that were not observed genome-wide.

The axon gene-set, a cellular component that conducts electrical signals to presynaptic boutons that store and release neurotransmitters, has also been found to play an important role in schizophrenia. Specifically, disruption in axon guidance and axonal growth have been associated with increased schizophrenia risk in both human and mouse models (48,49). A recent study showed that individuals at high genetic risk for schizophrenia had hemispherical asymmetry in whole-brain structural networks, indicating that genetic susceptibility to schizophrenia modulated white matter network abnormalities. Additionally, gene-set enrichment analysis found that genes participating in the PRS threshold used were involved in multiple relevant pathways, including axonal growth and axon guidance (18). The axon gene-set is therefore a highly relevant route to investigate in the context of schizophrenia. Here, the axon gene-set was associated with thalamic radiations, white matter microstructural tracts that link the thalamus to the rest of the cerebral cortex (50), and volume of the parahippocampal gyrus, a cortical region that plays a role in memory processes such as encoding and retrieval (51). Both neuroimaging phenotypes have been implicated in schizophrenia (52), and have recently been associated with state anhedonia and PRS for anhedonia, a core negative symptom of schizophrenia (53). The findings here indicate that genes conferring risk for schizophrenia that are aggregated in the axon gene-set are strengthening evidence for brain structural regions that are already implicated in schizophrenia psychopathology.

Finally, in this study the histone H3-K4 methylation gene-set was associated with entorhinal surface area. Histone H3-K4 methylation involves the modification of histone H3 by the addition of one or more methyl groups to lysine at position 4 of the histone. Histones, and specifically H3, are used to package DNA, and modifications lead to changes in gene expression (54). Epigenetic processes, including histone and DNA methylation, have been associated with schizophrenia through candidate gene regulation (*HDAC1, GAD67*) as well as epigenome-wide studies, lending support to the investigation of epigenetic modifications in schizophrenia (55–57). A key feature of schizophrenia is that it has its age of onset in young adults and the transcriptional regulation of schizophrenia risk genes encoding synaptic proteins occurs at the age of onset, potentially through mechanisms involving H3-K4 methylation (42).

The entorhinal cortex plays a role in the integration of multisensory information between cortical and subcortical structures. It is strongly connected to the hippocampus, and is involved in memory processes such as formation and consolidation (58). In studies investigating a neurodevelopmental animal model of schizophrenia, lesions in the entorhinal cortex, created early in the developmental period, were associated with enhanced mesolimbic dopamine release at a later timepoint, expressed through increased locomotor activity. This indicates that structural disruptions in this area are relevant for schizophrenia-like presentations (59). Interestingly, in an animal model, H3-K4 methylation was found to be upregulated in the hippocampus 1 hour after inducing contextual fear conditioning, a memory-related task which aims to rapidly create context-related fear memories (17). Although not directly related to the entorhinal cortex, this finding indicates that H3-K4 methylation may be a useful candidate in the investigation of some schizophrenia-related symptoms in a subcortical structure closely linked to the entorhinal region. Lastly, in a recent human study investigating cortical thickness, surface area, and folding index of the entorhinal cortex, Schultz et al. (2009) uncovered a link between left surface area and folding index and increased psychotic symptom severity, further supporting the role of the region in schizophrenia (58).

Interestingly, the dendritic spine and PSM gene-sets were not associated with any phenotypes investigated here. Both cellular components have been linked to schizophrenia previously and are closely related to the other biologically relevant pathways investigated here (i.e. PSD, axon gene-sets) (60,61). Results here may indicate that, while these gene-sets are still relevant in schizophrenia, we may need additional information (e.g. interaction with other relevant biological pathways or investigation of copy number variants in relation to these cellular components (62)) to identify their association with the clinical and structural brain features investigated here.

All psychotic-like experiences, as well as distress, were significantly associated with whole-genome PRS, as expected. This would indicate that the PRS generated from the schizophrenia GWAS are able to predict schizophrenia-related traits and are therefore a valuable tool here. In addition, a number of neuroimaging phenotypes were associated with whole-genome PRS (irrespective of which gene-set-specific SNPs were removed prior to calculation), including white matter microstructure: fractional anisotropy (global metrics, association fibres, thalamic radiations, cingulate gyrus, anterior and posterior thalamic radiations, and inferior longitudinal and inferior fronto-occipital fasciculi) and mean diffusivity (global metrics, association and projection fibres, thalamic radiations, corticospinal tract, anterior, superior, and posterior thalamic radiations, inferior longitudinal fasciculus, and cingulate gyrus); subcortical volumes: thalamus and accumbens; cortical regions: volume of the medial orbitofrontal cortex and superior temporal gyrus, and surface area of superior temporal gyrus. These areas have been associated with whole-genome PRS in previous studies, calculated both with the GWAS summary statistics utilised here, and with older GWAS (63,64). The findings here confirm previous results and provide additional evidence of an association between increased genetic risk for schizophrenia and disruptions in both grey and white matter.

There are a number of strengths in the current study. We utilised the largest, most recent GWAS of schizophrenia to determine associations in a large sample comprising clinical and neuroimaging datasets. Our findings were further tested utilising permutation analysis, indicating that our results were not due to chance. Finally, we were able to identify associations that could in future be utilised to identify stratified patient populations, potentially leading to earlier diagnosis and applied interventions.

Limitations of the current study include the investigation of these associations in a general population sample. This may partially explain low effect sizes observed throughout, and it is expected that these effect sizes will increase as clinical samples with available genetic and neuroimaging data increase. In addition, UK Biobank participants are generally healthier and wealthier than the general population, as shown in Fry et al. (2017) (65). Secondly, we are unable to generalise these findings to populations of non-European ancestry, as both GWAS and target samples were selected based on individuals of European descent. However, efforts are continually being made to collect data in other ancestries, and the associations identified here could be explored in these cohorts. A further limitation includes the use of the MHQ, which reports lifetime occurrence of PLEs, and not a specific timepoint. Therefore, our results here may differ in other age populations. Further, the questions asked are self-reported and it is unclear whether the experiences reported arose from normal experiences or were linked to a psychiatric disorder. However, the strong associations with schizophrenia whole-genome PRS indicate that the items pick up on some psychosis-related features.

The pathways investigated here were previously found to be implicated in schizophrenia in both animal and human studies (4,17,53). In the current study, we were able to identify structural neuroimaging as well as clinical phenotypes associated with biologically informed PRSs, and these were shown to be more strongly associated with neuroimaging phenotypes than whole-genome PRS. These findings indicate that genetic risk aggregated to biologically relevant pathways could be more informative than genome-wide risk and may be of relevance to future studies attempting to address heterogeneity and stratify individuals by genetic risk.

## Data Availability

All data produced in the present study are available upon reasonable request to the authors.

## Funding

This research was funded in whole, or in part, by the Wellcome Trust [216767/Z/19/Z]. This analysis uses linked data from UKB applications 4844 and 16124. For the purpose of open access, the author has applied a CC BY public copyright licence to any Author Accepted Manuscript version arising from this submission. MCB is supported by a Guarantors of Brain Non-clinical Post-Doctoral Fellowship and British Medical Association Margaret Temple Grant. HCW is supported by the Royal College of Physicians of Edinburgh (Sim Fellowship). MJA is supported by MRC Mental Health Data Pathfinder Award (MC_PC_17209). KM is supported by the Chief Scientist Office for Scotland and NHS Education for Scotland (PCL/17/01). This study is supported by a Wellcome Trust Strategic Award “Stratifying Resilience and Depression Longitudinally” (STRADL) (Reference 104036/Z/14/Z).

## Financial disclosures

The authors declare no conflicts of interest in relation to the work presented here. AMM has previously received funding from commercial sources (Pfizer, Roche, Abbvie, Sunovion, Jannssen and Lilly), but none of these funds or funders were used or involved in the current study.

## Acknowledgements

The authors wish to thank all the participants who are part of the UK Biobank cohort, and all of the UK Biobank staff who make this work possible. We would also like to thank our funders, Wellcome Trust, the Royal College of Physicians of Edinburgh, the Medical Research Council, the Chief Scientist Office for Scotland, NHS Education for Scotland, the Guarantors of Brain, and the British Medical Association.

## Data sharing statement

Data used in the preparation of this article were obtained from the UK Biobank cohort (https://www.ukbiobank.ac.uk/).

Scripts for the analyses in this project can be accessed at the following GitHub repository: https://github.com/mirunabarbu8/Fellowship-project.

## Notes

### Competing Interest Statement

The authors have declared no competing interest.

### Author Declarations

UK Biobank received ethical approval from the research ethics committee (REC reference 11/NW/0382), and this study has been approved by the UK Biobank Access Committee (Project No. 4844 and 16124).

### Summary of Updates

Include additional information

