## Supplementary Materials for "Pathway-based polygenic risk scores for schizophrenia and associations with clinical and neuroimaging phenotypes in UK Biobank"

Biological pathway information

The axon pathway represents the part of the neuron that conducts nerve impulses away from the cell bodies; electrical signals are then received by other neurons. The pathway is classified as a cellular component on the Gene Ontology database (GO term: 0030424 <http://amigo.geneontology.org/amigo/term/GO:0030424>).

The histone H3-K4 methylation pathway involves the modification of the H3 histone by addition of one or more methyl groups to lysine at position 4 of the histone. The pathway is classified as a biological process in Gene Ontology (GO term: 0051568; <http://amigo.geneontology.org/amigo/term/GO:0051568>). The pathway plays a role in brain development and cell differentiation. Dysregulated H3-K4 methylation has been associated with schizophrenia and autism (1).

The dendritic spine pathway is classified as a cellular component, comprises a membranous protrusion from dendrites that receives input from a presynapse, and can have variable spine morphology (GO term: 0043197; <http://amigo.geneontology.org/amigo/term/GO:0043197>). Dendritic spine impairments are present in schizophrenia (2).

The postsynaptic density pathway is a cellular component comprising an electron-dense protein network that is situated within and adjacent to the postsynaptic membrane of an asymmetric synapse (GO term: 0014069; <http://amigo.geneontology.org/amigo/term/GO:0014069>). The postsynaptic membrane comprises a specialised area of the membrane localised to the nerve ending and separated by the synaptic cleft; neurotransmitters cross this and transmit signals to the postsynaptic membrane (GO term: 0045211; <http://amigo.geneontology.org/amigo/term/GO:0045211>). Both have been previously implicated in schizophrenia, in genomic- and proteomic-specific studies (3).

|  | **Postsynaptic density** | **Postsynaptic membrane** | **Dendritic spine** | **Histone He-K4 methylation** | **Axon** |
| --- | --- | --- | --- | --- | --- |
| **N pathway genes in Trubetskoy et al. (2022)** (4) | 312/332 | 264/298 | 178/185 | 56/62 | 613/653 |
| **N SNPs** | 180,704 | 181,555 | 84,601 | 10,732 | 257,383 |

**Supplementary Table 1.** The number of genes in each pathway and those that are included in the discovery genome-wide association study (GWAS); the number of SNPs in each pathway that are present in the discovery GWAS.

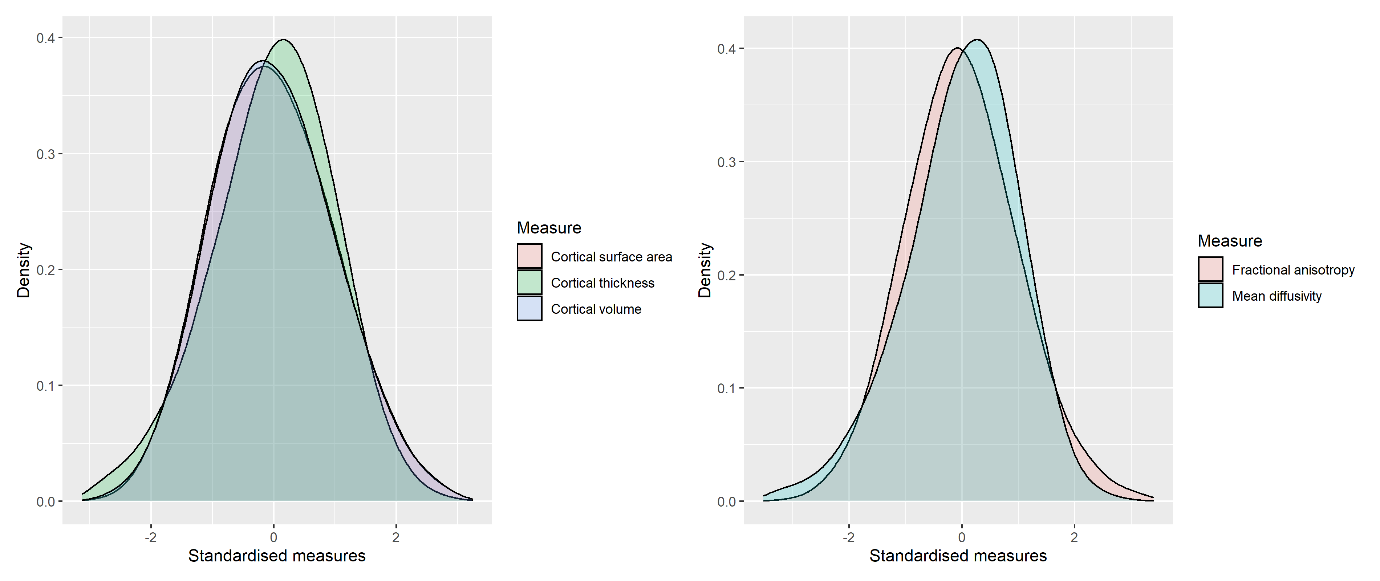

**Supplementary Figure 1**. Density plots of global measures of cortical regions (surface area, thickness, volume) and white matter microstructure (fractional anisotropy, mean diffusivity). The x-axis indicates standardised measures for each neuroimaging phenotype, and the y-axis represents the distribution density.

| **Imaging modality** | **Neuroimaging phenotype** |
| --- | --- |
| **Cortical regions (all regions have thickness, surface area, and volumes)** | **Global** |
|  | **Frontal lobe** |
|  | Superior frontal gyrus |
|  | Rostral middle frontal |
|  | Caudal middle frontal |
|  | Pars orbitalis |
|  | Pars triangularis |
|  | Pars opercularis |
|  | Lateral orbitofrontal |
|  | Medial orbitofrontal |
|  | Precentral gyrus |
|  | Paracentral cortex |
|  | **Temporal lobe** |
|  | Insula |
|  | Superior temporal |
|  | Transverse temporal |
|  | Middle temporal gyrus |
|  | Inferior temporal gyrus |
|  | Fusiform |
|  | Parahippocampal |
|  | Entorhinal |
|  | **Parietal lobe** |
|  | Postcentral gyrus |
|  | Paracentral cortex |
|  | Superior parietal cortex |
|  | Inferior parietal cortex |
|  | Supramarginal gyrus |
|  | Precuneus |
|  | **Occipital lobe** |
|  | Lateral occipital cortex |
|  | Cuneus |
|  | Pericalcarine cortex |
|  | Lingual gyrus |
|  | **Cingulate lobe** |
|  | Rostral anterior cingulate |
|  | Caudal anterior cingulate |
|  | Posterior cingulate cortex |
|  | Lingual gyrus |
| **DTI measures (FA and MD)** | **Global measure (PCA)** |
|  | **Projection fibres** |
|  | Forceps major |
|  | Forceps minor |
|  | Corticospinal tract |
|  | Acoustic radiation |
|  | Medial lemniscus |
|  | Middle cerebellar peduncle |
|  | **Association fibres** |
|  | Inferior fronto-occipital fasciculus |
|  | Uncinate fasciculus |
|  | Cingulum bundle (gyrus) |
|  | Cingulum bundle (parahippocampal) |
|  | Superior longitudinal fasciculus |
|  | Inferior longitudinal fasciculus |
|  | **Thalamic radiations** |
|  | Superior thalamic radiations |
|  | Posterior thalamic radiations |
|  | Anterior thalamic radiations |
| **Subcortical volumes** | Thalamus |
|  | Caudate |
|  | Putamen |
|  | Pallidum |
|  | Hippocampus |
|  | Amygdala |
|  | Accumbens |
|  | Intracranial volume |

**Supplementary Table 2.** Imaging modalities investigated.
